# Development and Psychometric Evaluation of a Bilingual Instrument for Assessing Beliefs Affecting Health-Seeking Intentions in Cognitive Decline Among Latino Populations

**DOI:** 10.1101/2025.04.24.25326381

**Authors:** Maria Mora Pinzon, Philip Sayegh, Susana Fernandez de Cordova, Roger Brown, Bruce Barrett

## Abstract

**Background:** The Latino community in the United States is disproportionately affected by Alzheimer’s disease and related dementias (ADRD). However, there are no instruments to assess health seeking behaviors in this population. This study describes the development and validation process of a new 35-item instrument, “**Be**liefs affecting health **S**eeking **I**ntentions in **C**ognitive decline (BESIC)”, available in both English and Spanish.

**Methods:** The psychometric analysis involved assessing the goodness of fit of three measurement models; congeneric model, tau-equivalency, and the parallel model. Multigroup Confirmatory Factor Analysis (MGCFA) was used to check if the same factors are being measured in the same way across different groups.

**Results:** The partial tau-equivalent model provided the best fit for our data, suggesting that all items in the instrument measure the same underlying concept construct with different degrees of precision and error. The instrument demonstrated good reliability for all sub-domains in the total sample, as well as for the two language surveys. While average scores in the two language groups were somewhat different, MGCFA demonstrated that the instrument works well and similarly in both English and Spanish.

**Conclusion:** These psychometric validation findings suggest that BESIC is a useful tool for measuring beliefs affecting health-seeking intentions during cognitive decline in both English and Spanish-speaking populations in the United States. However, while the overall structure of the instrument was equivalent across languages, the strength of these relationships and the average scores on the items were not. This suggests that the way individuals from different language groups respond to the items may vary, requiring further investigation. This will help refine the instrument further and ensure its accuracy and usefulness in future research and practice.

## INTRODUCTION

Latino individuals are approximately 1.5 times more likely to develop Alzheimer’s disease and related dementias (ADRD) than non-Latino whites,^1^ and have higher rates of under-diagnosis.^2^ Reasons for higher rates of under-diagnosis include the commonly-held belief among Latino individuals that ADRD symptoms are part of normal aging,^3^ lack of ADRD knowledge,^4,5^ and limited access to culturally-safe competent/language appropriate healthcare services.^6^ Understanding the role that knowledge and beliefs have on health seeking behaviors is necessary to inform the development of interventions responsive to the unique needs of Latino communities, which are both diverse and changing as the proportion of Mexican-Americans decrease, and the proportion of US-born Latinos increases.^7–9^

Most of the existing validated instruments for ADRD were designed for healthcare professionals or English-speaking communities.^10^ Those that have been validated in Spanish explore only knowledge about ADRD and not beliefs or intentions, resulting in gaps in our understanding of how health seeking behaviors are modified by knowledge. One of the instruments currently available is a 26-item scale, the Cultural Beliefs about Alzheimer’s Disease (CBAD), which aims to measure how cultural beliefs influence healthcare-seeking behaviors. Preliminary use of CBAD in two studies revealed that many Mexicans and Mexican Americans perceive Alzheimer’s disease as a normal part of aging or a mental illness caused by bad habits.^5,11^ However, CBAD has not yet undergone a validation study to assess its validity, reliability, and generalizability to Latino individuals from countries other than Mexico.

The purpose of this study is to describe the development process of a new 35-item instrument, available in both English and Spanish, titled “**BE**liefs affecting health **S**eeking **I**ntentions in **C**ognitive decline (BESIC),” which seeks to explore knowledge and beliefs about ADRD, health seeking behavior intentions, and stigma in Latino communities. This paper will also detail the psychometric characteristics of BESIC. This instrument can be used to understand the prevalence of attitudes, beliefs, knowledge, and intention to seek care within each community, which in turn can be used to develop interventions that increase access to care.

## METHODS

The study underwent an ethical review by the Institutional Review Board (IRB) at the University of Wisconsin - Madison, and was determined to meet the criteria for exempt human subjects research as defined under 45 CFR 46: Category 2: Research involving the use of educational tests, surveys, interviews. This study aligns with the STROBE guidelines^12^ for observational studies, ensuring rigorous methodology and robust findings.

### Theoretical Framework

We used the theory of planned behavior (TPB) to build the instrument.^13^ This theoretical framework posits that behaviors are influenced by three primary domains: 1) Behavioral beliefs, which pertain to the perceived benefits associated with a behavior; 2) Normative beliefs, which relate to the social acceptability of the behavior; and 3) Control beliefs, which refer to the perceived ease or difficulty of performing the behavior.

Research has demonstrated that knowledge and attitudes can significantly impact these beliefs.^14^ For instance, increased knowledge about a particular behavior can enhance one’s behavioral beliefs, while positive attitudes can strengthen normative and control beliefs. Therefore, it was essential to consider these additional factors when developing a comprehensive measurement instrument. For this study, the behavior of interest is seeking care for memory problems that affect daily living. The instrument was specifically designed to explore the domains of the TPB, incorporating measures of knowledge and attitudes to provide a more holistic understanding of the factors influencing this behavior.

### Instrument Development

In a previous study (not published), we explored the cultural and psychological factors that influence health-seeking behaviors for ADRD in Latino individuals through structured interviews with 24 Latino adults older than 50 years old. We used the interview responses to modify the *Cultural Beliefs about ADRD (CBAD)* survey,^5^ and *the Attitudes of People from Ethnic Minorities to Help-Seeking for Dementia (APEND) instrument*,^13^ which was developed for Southeast Asian communities in the UK.^8^ Cultural and linguistic adaptation was needed to ensure the use of common cultural elements unique to Latino individuals living in the US. Additionally, we included modified items from the validated-instrument Basic Knowledge in ADRD scale^15^ to explore the domains related to knowledge. The modifications included changing from True/False to a Likert Scale and deleting some items that were repetitive with other items from the CBAD survey.

The resulting instrument is titled “***Beliefs affecting health-seeking intentions in cognitive decline (BESIC)***, which contains 35-items using that are rated on a 5-point Likert-Scale (Supplement 1) and is available in English and Spanish. For the Spanish version of the instrument, the linguistic translation was done by a professional translator, and then reviewed by three native-Spanish speakers from Puerto Rico, Peru, and Venezuela, who performed successive iterative revisions of the instrument. This method was used because the goal was culturally and linguistically adapt the questions from English rather than perform a literal translation, consistent with methodology used in other studies, including the translation of the Uniform Data Set of the NIA Alzheimer’s Disease Centers.^16^

### Cognitive Interviews

We conducted basic cognitive interviews to enhance the validity and reliability of our survey instrument. The purpose of cognitive testing was to assess participants’ comprehension, interpretation, and reactions to survey questions, ensuring that they align with the intended meaning and capture respondents’ perspectives accurately.^17^ We recruited 10 individuals from of Latino origin living in Wisconsin (Countries of Origin: Mexico, Colombia, El Salvador, and Nicaragua). These participants received the survey in both English and Spanish and were asked to complete the survey in the language of their choice. A telephone survey interviewer from the University of Wisconsin Survey Center, who was a native Spanish speaker, conducted short interviews with each participant. During these interviews, participants were asked about their reactions to survey questions, the wording, and their overall thoughts about the survey. UWSC collected interview responses and interviewer notes for subsequent review. Each participant who completed the interview received $50. The insights gained from cognitive testing informed revisions to the survey instrument, ensuring its suitability for the target population and research objectives.

### Survey Recruitment via Mail

The target population for this study was Latino adults 18 years and over that live in Milwaukee and Dane Counties in Wisconsin. Between July 2021, and Sept 2021, UWSC mailed surveys in both English and Spanish were sent to 750 households in Madison and Milwaukee. Addresses were drawn from an Address-Based Sample (ABS),^18^ which utilizes postal addresses that were purchased from Marketing Systems group. To maximize recruitment of the target group, simple random sampling was conducted within specific census block groups characterized by high Hispanic density in Milwaukee and Dane Counties.

Each survey package included an introductory letter from the Principal Investigator and a $5 incentive to encourage participation.^19^ A postcard reminder was sent after two (2) weeks, followed by a second survey to encourage participation from non-respondents.

The selection of the household member who would participate was left to the household to determine. However, the invitation letter noted that the best person to fill it out would be an adult 60 years of age or older, or an adult 18 or older who provides care for an older adult.

Response Rate was calculated by dividing the number of completed surveys by the number of mailed surveys minus the number undeliverable surveys. This method is described by the American Association for Public Opinion Research Transparency Initiative.^20^

### Survey Recruitment via in-person outreach

Between June 2022 – and June 2023, we employed targeted in-person outreach within the Hispanic/Latino community in Wisconsin to enhance survey participation. Collaborating with community organizations and participating in local events, we engaged with potential participants. Trained research staff or the Principal Investigator (PI) explained the survey’s purpose, emphasizing its relevance to health and dementia awareness. Potential participants were informed of the voluntary nature of participation. Participants could choose to complete the survey in either English or Spanish. Recognizing potential language barriers, we offered assistance to those who had difficulty reading the survey. Our research staff clarified questions and provided support throughout the process. No personal health information was collected. Individuals who completed the survey received a $15 gift card or a cash incentive.

In addition, we distributed study information flyers through community organizations, list-servers, and direct outreach in private or public spaces. These flyers included a QR code or a direct link to the study page. Participants could access the survey independently, without requiring assistance. After completing the survey, participants were prompted to provide their contact information to receive a digital gift card for $15 to the store of their choice.

### Data management

Completed surveys were entered into RedCap^21^ using a double entry method. This rigorous approach involved two independent research staff members entering the same survey data separately. The dual entries were then electronically compared by a data scientist using REDCap’s data comparison tool, which identified any discrepancies between the two sets of entries. When inconsistencies were detected, they were carefully reviewed by the data scientist who, when necessary, referred to the original survey for accurate resolution. This process reduced data entry errors, ensured high data quality, and provided an essential audit trail.

### Psychometric Analysis

*“The Beliefs affecting health-seeking intentions in cognitive decline*” (BESIC) scale was analyzed in three stages using classical test theory, focusing on the domains of knowledge-likelihood, knowledge-risk, knowledge-prevention, behavioral beliefs, and normative & control beliefs.

In stage 1, a confirmatory factor analysis model was constructed to evaluate the congeneric structure of the BESIC domains in both English and Spanish speaking samples (n=249). The model was estimated using weighted least square mean and variance adjusted (WLSMV) estimation, a probit link, and the theta parameterization.^22^ This allowed for an examination of the relationships between the observed variables and the latent factors.

Stage 2 focused on assessing the reliability of the BESIC domains. Coefficient alpha, a widely used measure of internal consistency reliability, was initially considered. However, since coefficient alpha may underestimate reliability if the assumption of tau-equivalent measurement models is not met, different measurement models were evaluated. These included the congeneric model, the tau-equivalent model, and the parallel model.^23^ The fit of these models was assessed to determine the most appropriate measure of reliability for the BESIC domains.

Stage 3 involved assessing the factorial invariance of the BESIC domains across the two language groups (English n = 119 and Spanish n = 130) using classical test theory. This was done to determine if the same underlying constructs were being measured in both groups. Four levels of factorial invariance were considered: configural invariance, weak invariance, strong invariance, and very strong invariance. Configural invariance assumes that the same pattern of factor loadings occurs across groups, while weak invariance assumes that the factor patterns are equal.

Strong invariance adds constraints on factor coefficients and intercepts, and very strong invariance constrains error terms to equality. Fit indices such as Tucker-Lewis index, Akaike information criterion, Bayesian information criterion, comparative fit index, model chi-square, ratio of chi-square to degrees of freedom, and chi-square difference test were used to assess the fit of the models.^24^ Achieving at least configural to weak invariance is necessary for the use of the BESIC instrument, while strong invariance allows for more meaningful comparisons of latent means across groups. These calculations were performed using the statistical software Mplus Version 8.11.^25^

## RESULTS

### Recruitment Results

A total of 353 surveys were completed across the different data collection methods, with 283 (80%) self-identifying as Hispanic/Latino, 70% female, and 52% of them had some college education or completed college. Table 1 shows additional demographic characteristics of the respondents according to method of collection (Online vs. Paper vs Mail).

**Table 1.** Demographics of survey respondents according to recruitment method.

|  |  | Direct Outreach |  |  |
| --- | --- | --- | --- | --- |
|  | Mail (n=117) | Online Responses (n=149) | Paper Responses (n=87) | Total (n=353) |
| **Age** |  |  |  |  |
| 18 – 25 years | 7 (6%) | 77 (52%) | 17 (20%) | 101 (28.61%) |
| 26 – 64 years | 81 (69%) | 57 (38%) | 54 (62%) | 192 (54.39%) |
| => 65 years | 29 (25%) | 15 (10%) | 16 (18%) | 60 (17.00%) |
| **Gender** |  |  |  |  |
| Man | 24 (21%) | 44 (31%) | 22 (26%) | 90 (25.50%) |
| Non-Binary | - | 2 (1%) | 1 (1%) | 3 (0.85%) |
| Woman | 91 (78%) | 96 (68%) | 62 (73%) | 249 (70.54%) |
| **Ethnicity** |  |  |  |  |
| Hispanic | 51 (46%) | 147 (100%) | 85 (100%) | 283 (80.17%) |
| Not Hispanic | 59 (54%) | - | - | 59 (16.71%) |
| **Country or Region of Origin or Heritage** |  |  |  |  |
| Centro America | - | 8 (7%) | 3 (4%) | 11 (3.12%) |
| Mexico | 35 (69%) | 56 (50%) | 46 (66%) | 137 (38.81%) |
| Puerto Rico and Caribbean | 15 (30%) | 8 (7.25%) | 5 (7%) | 28 (7.93%) |
| South America | 1 (2%) | 40 (36%) | 16 (22%) | 57 (16.15%) |
| **Education** |  |  |  |  |
| No formal education | 1 (1%) | 2 (1%) | 4 (5%) | 7 (1.98%) |
| Elementary School | 8 (7%) | 1 (1%) | 11 (12%) | 20 (5.67%) |
| Middle School | 3 (3%) | 4 (3%) | 4 (5%) | 11 (3.12%) |
| High School | 47 (40%) | 27 (18%) | 19 (22%) | 93 (26.35%) |
| Technical or Trade School | 11 (9%) | 10 (7%) | 4 (5%) | 25 (7.08%) |
| Some College | 23 (20%) | 45 (30%) | 7 (8%) | 75 (21.25%) |
| College or more | 23 (20%) | 53 (36%) | 34 (38%) | 110 (31.16%) |

Column Percentages

Regarding the survey mailed to households, out of the 750 surveys mailed, the total number of completed questionnaires was 117, 15 were undeliverable, and 19 were vacant households. The overall response rate for the survey was 15.6%. Of these, 51 responses (47%) self-identified/Hispanic or Latino (Table 1). Because the other data collection methods do not have a defined denominator, response rate could not be provided.

Online recruitment was particularly effective in engaging younger respondents (18-25 years), whereas mail and paper methods captured a more balanced age distribution, including a higher proportion of older adults (26-64 and 65+ years). Similarly, significant number of respondents with higher education levels (college or more) participated via online and paper responses, and there was a notable presence of respondents with no formal education or only elementary school education, particularly in paper responses (5% and 12%, respectively).

### Descriptive Statistics

Missing data were assessed and cases that had less than 70% complete data were initially removed, which left a sample of 249 cases. These remaining 249 cases were assessed for meeting the assumption of Missing Completely at Random (MCAR)^26^ or Missing at Random (MAR). Half of the sub-domains met the assumption of MCAR, based on Little’s test,^26^ the other half met the assumption of MAR.^26^ These missing values were then imputed using multiple imputation by chained equations providing a final analyzable sample size of 249 (English = 119, Spanish = 130).^27^ Item correlations are provided in **Supplemental File 3** as polychoric correlations, due to the categorical nature of the items.

#### Stage 1

The least restrictive five factor *congeneric* measurement model is provided in Table 2, for each of the five domains. Although the five-domain structure did not achieved tau-equivalency, all domains did achieve partial tau-equivalency. Table 3 provides the final domain loadings for the congeneric BESIC scale.

**Table 2.** Goodness of fit for four measurement models and bifactor model of the BESIC Scale.

| Model | Phase | $\chi^2$<br>(df) | p-value | $\chi^2$/df<br>ratio | CFI | TLI | $\Delta$ CFI | $\Delta$ TLI | $\chi^2_{diff}$<br>(df) | p-value | Decision |
| --- | --- | --- | --- | --- | --- | --- | --- | --- | --- | --- | --- |
| Model 1a: 5 Factor Congeneric Model | 1 | 1196.6<br>(584) | <0.001 | 2.04 | 0.919 | 0.913 |  |  | --- | --- | Reject.<br>Not all loadings were significant |
| Model 1b: 5 Factor congeneric Model less item 9c | 1 | 1178.6<br>(550) | <0.001 | 2.14 | 0.917 | 0.910 | -0.002 | -0.003 |  |  | Accept |
| Model 2: 5 Factor tau-equivalent Model | 2 | 1574.6<br>(580) | <0.001 | 2.71 | 0.869 | 0.865 | -0.048 | -0.045 |  |  |  |
|  |  |  |  |  |  |  |  |  | 328.8<br>(30) | <0.001 | Reject |
| Model 3: 5 Factor partial tau-equivalent model: | 2 | 1165.4<br>(565) | <0.001 | 2.06 | 0.921 | 0.916 | +0.052 | +0.051 |  |  |  |
|  |  |  |  |  |  |  |  |  | 20.8<br>(15) | 0.14 | Accept |

Model 1a was rejected due to a single item (Item 9c) not loading.

**Table 3.** Final standardized loadings for the five factor congeneric model.

| **Domain Knowledge - Likelihood** | Parameter loading | Standard error | Z-value | p-value |
| --- | --- | --- | --- | --- |
| How <u>likely</u> is someone to have Alzheimer's disease if... |  |  |  |  |
| ...they stop taking part in social activities? | 0.410 | 0.053 | 7.682 | 0.000 |
| ...they wear a heavy coat when it is hot outside? | 0.461 | 0.052 | 8.902 | 0.000 |
| ...they forget names of familiar objects? | 0.724 | 0.030 | 24.522 | 0.000 |
| ...they often forget appointments? | 0.851 | 0.020 | 42.223 | 0.000 |
| ...they have difficulty remembering the rules of a game they have played many times before? | 0.836 | 0.020 | 42.014 | 0.000 |
| ...they have trouble following directions? | 0.853 | 0.018 | 47.352 | 0.000 |
| ...they ask someone the same question over and over again? | 0.846 | 0.021 | 39.963 | 0.000 |
| ...they have trouble counting money? | 0.818 | 0.022 | 36.837 | 0.000 |
| ...they lose their keys from time to time? | 0.776 | 0.026 | 29.585 | 0.000 |
| **Model fit: chi-square (df) = 129.5 (27) p <0.001, CFI = 0.971, TLI = 0.962 SRMR = 0.041** |  |  |  |  |
| **Domain Knowledge - Risk** |  |  |  |  |
| How much is someone's risk of getting Alzheimer's disease <u>increased</u> if... |  |  |  |  |
| ...a parent had it? | 0.341 | 0.055 | 6.242 | 0.000 |
| ...they have a serious hit to their head? | 0.510 | 0.043 | 11.767 | 0.000 |
| ...they have high blood sugar? | 0.820 | 0.025 | 33.127 | 0.000 |
| ...they have high blood pressure? | 0.818 | 0.025 | 33.241 | 0.000 |
| ...they have had a stroke? | 0.710 | 0.031 | 22.798 | 0.000 |
| ...they have poor nutrition? | 0.784 | 0.025 | 1.649 | 0.000 |
| ...they do not get enough sleep? | 0.813 | 0.025 | 32.030 | 0.000 |
| ...they do not take care of themselves? | 0.659 | 0.035 | 18.918 | 0.000 |
| ...they experienced a difficult life or traumatic event? | 0.610 | 0.039 | 15.641 | 0.000 |
| **Model fit: chi-square (df) = 353.1 (27) p <0.001, CFI = 0.870, TLI = 0.826 SRMR = 0.094** |  |  |  |  |
| **Domain Knowledge - Prevention** |  |  |  |  |
| To prevent or slow Alzheimer's disease, how <u>important</u> is it to... |  |  |  |  |
| ...get help with dementia symptoms early? | 0.777 | 0.031 | 24.837 | 0.000 |
| ...stay mentally active? | 0.805 | 0.029 | 28.210 | 0.000 |
| ...be with others to keep the memory sharp? | 0.783 | 0.028 | 27.833 | 0.000 |
| ...have your memory tested as part of your regular check-up if you are over 65? | 0.708 | 0.033 | 21.430 | 0.000 |
| ...stay physically active? | 0.705 | 0.031 | 22.769 | 0.000 |
| ...keep written lists as reminders? | 0.692 | 0.035 | 19.889 | 0.000 |
| ...try very hard to remember things? | 0.608 | 0.039 | 15.419 | 0.000 |
| ...take care of high blood pressure? | 0.586 | 0.042 | 13.940 | 0.000 |
| **Model fit: chi-square (df) = 188.0 (20) p =0.001, CFI = 0.919, TLI = 0.887 SRMR = 0.074** |  |  |  |  |
| **Domain – Behavioral Beliefs** |  |  |  |  |
| If you were having memory problems that affected your daily routine... |  |  |  |  |
| ...how <u>likely</u> would you be to go see a doctor? | 0.680 | 0.040 | 17.026 | 0.000 |
| ...how <u>confident</u> are you that doctors would be able to tell the cause of memory problems? | 0.761 | 0.031 | 24.490 | 0.000 |
| ...how <u>important</u> would it be to know the cause of the memory problems? | 0.591 | 0.046 | 12.945 | 0.000 |
| ...how <u>confident</u> are you that doctors would be able to provide treatment to help with memory problems? | 0.833 | 0.031 | 27.305 | 0.000 |
| ...how <u>confident</u> are you that doctors would be able to teach your family how to take care of you? | 0.753 | 0.030 | 25.347 | 0.000 |
| **Model fit: chi-square (df) = 44.2 (5) p =<0.001, CFI = 0.965, TLI = 0.930 SRMR = 0.041** |  |  |  |  |
| **Domain – Normative & Control Beliefs** |  |  |  |  |
| If you were having memory problems that affected your daily routine... |  |  |  |  |
| ...how <u>likely</u> would it be that your family would approve of you seeking help from a doctor? | 0.884 | 0.035 | 25.058 | 0.000 |
| ...how <u>likely</u> is your family to think that you should go to the doctor? | 0.893 | 0.037 | 24.419 | 0.000 |
| ...how <u>easy</u> would you find it to seek help from a doctor for memory problems? | 0.554 | 0.047 | 11.679 | 0.000 |
| ...how <u>useful</u> would it be to seek help from a doctor for memory problems? | 0.628 | 0.041 | 15.381 | 0.000 |
| **Model fit: chi-square (df) = 48.3 (2) p =<0.001, CFI = 0.963, TLI = 0.888 SRMR = 0.047** |  |  |  |  |

#### Stage 2

We then evaluated the reliability of each sub-domain using coefficient alpha and McDonald’s omega coefficient.^28^ Omega coefficient assesses the proportion of variance in domain scores attributed to a sub-domain, and when the tau-equivalency assumption is met, coefficient alpha and omega are equivalent. Table 4 provides reliability coefficients for the entire sample, and each sub-sample (English speaking and Spanish speaking). Results reveal that coefficient alpha was lower than expected due to partial acceptance of the tau-equivalency assumption. Thus, Omega coefficient provided a more accurate measure of reliability than coefficient alpha. All sub-domains had suitable internal consistency. The reliability coefficients are a measure of internal consistency of a scale or measure, ranging from 0 to 1. A coefficient of 1 is considered perfect reliability, 0.9 or higher is considered excellent, 0.8-0.9 is good, 0.7-0.8 is acceptable, 0.6-0.7 is questionable, 0.5-0.6 is poor, and below 0.5 is unacceptable.

**Table 4.** Reliability of the domains.

| Domains | Cronbach Alpha | Omega Coefficients |
| --- | --- | --- |
| Full Sample |  |  |
| Knowledge-Likelihood | 0.884 | 0.908 |
| Knowledge-Risk | 0.805 | 0.816 |
| Knowledge-Prevention | 0.824 | 0.891 |
| Behavioral Beliefs | 0.768 | 0.844 |
| Normative & Control Belief | 0.760 | 0.801 |
| English Language |  |  |
| Knowledge-Likelihood | 0.882 | 0.907 |
| Knowledge-Risk | 0.848 | 0.861 |
| Knowledge-Prevention | 0.847 | 0.922 |
| Behavioral Beliefs | 0.814 | 0.895 |
| Normative & Control Belief | 0.760 | 0.838 |
| Spanish Language |  |  |
| Knowledge-Likelihood | 0.888 | 0.910 |
| Knowledge-Risk | 0.836 | 0.844 |
| Knowledge-Prevention | 0.832 | 0.889 |
| Behavioral Beliefs | 0.799 | 0.880 |
| Normative & Control Belief | 0.760 | 0.823 |

#### Stage 3

Factorial invariance (Table 5) was assessed on the five-factor congeneric model across two language groups (English speaking and Spanish speaking) using classical test theory. The invariance included configural invariance, weak invariance, strong invariance, and very strong invariance. Generally, both configural and weak invariance was supported in all five domains, with configural accepted in all domains. To consider the use of the BESIC scale for both English and Spanish speakers, at least configural to weak factorial invariance should be achieved. This means that the same pattern of zero and nonzero factor loadings is assumed to occur across groups. Additionally, the factor coefficients and intercepts should be constrained to be equal across groups. The ideal scenario is for strong factorial invariance to be achieved, as this supports the most meaningful interpretation of latent means across groups.

**Table 5.** Language Invariance Analysis.

| Model | $\chi^2$<br>(df) | p-value | $\chi^2$/df<br>ratio | English<br>$\chi^2$<br>(%) | Spanish<br>$\chi^2$<br>(%) | CFI | TLI | RMSEA | SRMR | $\chi^2_{diff}$<br>(df) | p-value | Decision |
| --- | --- | --- | --- | --- | --- | --- | --- | --- | --- | --- | --- | --- |
| Configural Invariance<br>5 Domains | 1819.3<br>(1200) | <0.001 | 1.51 | 919.9<br>(50.5%) | 899.4<br>(49.5%) | 0.917 | 0.917 | 0.064 | 0.091 |  |  | Accept |
| Individual Domains |  |  |  |  |  |  |  |  |  |  |  |  |
| Domain 1<br>Knowledge-Likelihood | 191.8<br>(80) | <0.001 | 2.39 | 61.2<br>(31.9%) | 130.6<br>(68.1%) | 0.972 | 0.974 | 0.106 | 0.053 |  |  | Accept |
| Domain 2<br>Knowledge-Risk | 390.2<br>(80) | <0.001 | 4.87 | 182.6<br>(46.7%) | 207.6<br>(53.3%) | 0.876 | 0.889 | 0.176 | 0.098 |  |  | Reject |
| Domain 3<br>Knowledge-Prevention | 248.3<br>(63) | <0.001 | 3.94 | 139.0<br>(56.0%) | 109.2<br>(44.0%) | 0.919 | 0.928 | 0.154 | 0.077 |  |  | Accept |
| Domain 4<br>Behavioral Beliefs | 81.7<br>(24) | <0.001 | 3.40 | 35.2<br>(43.0%) | 46.5<br>(57.0%) | 0.946 | 0.955 | 0.139 | 0.052 |  |  | Accept |
| Domain 5 Normative &<br>Control | 49.5<br>(15) | <0.001 | 3.30 | 14.0<br>(28.2%) | 35.5<br>(71.8%) | 0.975 | 0.980 | 0.136 | 0.051 |  |  | Accept |
| Metric Invariance<br>5 Domains | 1921.9<br>(1235) | <0.001 | 1.55 | 1038.6<br>(54.0%) | 883.3<br>(46.0%) | 0.907 | 0.911 | 0.067 | 0.093 | 125.1<br>(35) | <0.001 | Reject |
| Individual Domains |  |  |  |  |  |  |  |  |  |  |  |  |
| Domain 1<br>Knowledge-Likelihood | 190.7<br>(89) | <0.001 | 2.14 | 79.7<br>(41.8%) | 110.9<br>(58.2%) | 0.974 | 0.979 | 0.096 | 0.059 | 20.3<br>(9) | 0.015 | Reject |
| Domain 2<br>Knowledge-Risk | 462.2<br>(89) | <0.001 | 5.19 | 270.9<br>(58.6%) | 191.3<br>(41.4%) | 0.851 | 0.879 | 0.184 | 0.109 | 80.0<br>(9) | <0.001 | Reject |
| Domain 3<br>Knowledge-Prevention | 194.5<br>(71) | <0.001 | 2.73 | 109.2<br>(56.1%) | 85.3<br>(43.9%) | 0.946 | 0.957 | 0.118 | 0.076 | 7.3<br>(8) | 0.49 | Accept |
| Domain 4<br>Behavioral Beliefs | 95.6<br>(29) | <0.001 | 3.29 | 52.5<br>(54.9%) | 43.0<br>(45.1%) | 0.938 | 0.957 | 0.136 | 0.067 | 20.5<br>(5) | 0.001 | Reject |
| Domain 5 Normative &<br>Control | 59.4<br>(19) | <0.001 | 3.12 | 23.4<br>(39.3%) | 36.0<br>(60.7%) | 0.971 | 0.982 | 0.131 | 0.062 | 14.7<br>(4) | 0.005 | Reject |
| Scalar Invariance<br>5 Domains | 1923.4<br>(1270) | <0.001 | 1.51 | 1023.1<br>(53.1%) | 900.2<br>(46.9%) | 0.912 | 0.917 | 0.064 | 0.097 | 156.6<br>(70) | <0.001 | Reject |
| Individual Domains |  |  |  |  |  |  |  |  |  |  |  |  |
| Domain 1<br>Knowledge-Likelihood | 171.6<br>(98) | <0.001 | 1.75 | 75.9<br>(44.2%) | 95.6<br>(55.8%) | 0.981 | 0.986 | 0.078 | 0.063 | 29.0<br>(18) | 0.048 | Reject |
| Domain 2<br>Knowledge-Risk | 404.3<br>(98) | <0.001 | 4.12 | 220.5<br>(54.5%) | 183.7<br>(45.5%) | 0.878 | 0.910 | 0.158 | 0.116 | 83.3<br>(18) | <0.001 | Reject |
| Domain 3<br>Knowledge-Prevention | 187.5<br>(79) | <0.001 | 2.37 | 104.7<br>(55.8%) | 82.7<br>(44.2%) | 0.953 | 0.966 | 0.105 | 0.078 | 19.3<br>(16) | 0.25 | Accept |
| Domain 4<br>Behavioral Beliefs | 91.6<br>(34) | <0.001 | 2.69 | 51.1<br>(55.8%) | 40.4<br>(44.2%) | 0.947 | 0.969 | 0.117 | 0.065 | 24.7<br>(10) | 0.005 | Reject |
| Domain 5 Normative &<br>Control | 67.9<br>(23) | <0.001 | 2.95 | 30.2<br>(44.4%) | 37.7<br>(55.6%) | 0.967 | 0.983 | 0.125 | 0.062 | 24.5<br>(8) | 0.002 | Reject |

CFI: Comparative Fit Index; TLI: Tucker-Lewis Index; RMSEA: Root Mean Square Error of Approximation, SRMR: Standardized Root Mean Square Residual.

## DISCUSSION

The development and validation of a novel instrument is a multifaceted endeavor, requiring meticulous attention to detail. Our study introduces innovative elements that significantly contribute to the field. Specifically, we highlight three key aspects: diverse population recruitment, cultural appropriateness assessment, and bilingual psychometric validation.

Our study stands out for its deliberate efforts to recruit a diverse participant pool. By engaging individuals from various demographic backgrounds, including age, literacy levels, and cultural heritage, we ensured a representative sample. This inclusivity enhances the generalizability of our findings and underscores the instrument’s relevance across Latino communities. This approach is particularly crucial given the heterogeneity of Latino populations in the United States, which often goes unaddressed in health research.

The response rate of 15% on our survey distributed by mail is generally consistent with other literature on survey response rates among Latino populations (range 10 – 30%),^29,30^ although it falls on the lower end of the spectrum. It’s important to note that mailed surveys often yield lower response rates compared to other modes of data collection for Latino populations,^31,32^ requiring the use of multiple modalities.^33^ However, the groups engaged in each method of data collection will be different. A meta-analysis by Daikeler et al. found that online surveys had an average response rate of 12% lower than other survey modes, and this gap was even larger for populations with lower internet access or digital literacy, such as older adults.^34^ In contrast, younger generations have higher rates of internet usage and comfort with online platforms, making them more likely to participate in web-based surveys.

Beyond linguistic equivalence, we meticulously evaluated the cultural appropriateness of the instrument. Cultural nuances impact how individuals perceive and respond to survey items. Our rigorous process involved cognitive interviews, expert reviews, and pilot testing to ensure that the instrument resonated with the cultural context of our target population. The use of cognitive interview techniques enhances the validity of our measurements and follows best practices in the literature, as described by Hawkins et al. who emphasized the need for qualitative methods, including descriptions of the intent for each item/question, to ensure construct equivalence across languages.^35^

Regarding our psychometric analyses, we found that the partial tau-equivalent model was the most appropriate measure of reliability for this data set. This means that while all the items in our instrument are measuring the same underlying concept, they do so with different degrees of precision and error. This is an important finding as it suggests that our instrument is capturing a range of nuances within this conceptual entity, rather than just one aspect of it. This is consistent with previous studies that have also found the partial tau-equivalent model to be a more accurate measure of reliability when the assumption of tau-equivalence is not met.^23^

A groundbreaking feature of our research lies in the concurrent validation of the instrument in both English and Spanish. Few studies have attempted such dual-language validation. By doing so, we address the linguistic diversity within Latino populations and provide a robust tool that transcends language barriers. Our approach aligns with best practices for cross-cultural research and ensures that the instrument captures the intended constructs accurately, regardless of language preference.

Our analysis showed that the instrument works similarly well in both English and Spanish. This was assessed using Multigroup Confirmatory Factor Analysis (MGCFA), which checks if the same factors (or underlying concepts) are being measured in the same way across different groups. In our case, these groups were English and Spanish speakers. This is crucial because it means that our instrument is not biased towards one language group and can be used reliably across both. This finding aligns with the principles of measurement invariance, which state that the observed scores should depend only on latent construct scores, and not on group membership.^36^

However, it is important to note that while the overall structure of the instrument (i.e., the pattern of factor loadings or how each item relates to the underlying concept it is supposed to measure) was equivalent across languages (configural invariance), the strength of these relationships (metric invariance) and the average scores on the items (scalar invariance) were not. This suggests that while the instrument is measuring the same concepts across languages, the way individuals from different language groups respond to the items may vary. These results attest to the instrument’s stability and reliability, positioning it as a valuable tool for future investigations.

There are limitations to this study, including its survey-based nature. Specifically, our recruitment efforts were focused on Wisconsin, which, although diverse in reaching various demographic groups, may not fully represent the entire diversity within the Hispanic/Latino community across the United States. Additionally, the translation and adaptation process of the survey instrument introduces variability that could impact the responses. While we diligently reviewed the survey with native speakers, nuances of language and culture might not have been fully captured. For instance, after the survey was completed, individuals from the Caribbean pointed out that the term used to describe high school in some South American countries *(“Bachillerato”)* was associated with complete college education in their country—a nuance that was not identified during our earlier stages. Furthermore, our approach to handling missing data, although rigorous, may still influence the results.

## CONCLUSION

Overall, we found that the BESIC instrument had good reliability for all sub-domains across the total sample, as well as for the two separate English- and Spanish-language subsamples. This suggests that our instrument is a reliable way of assessing beliefs influencing health-seeking intentions in cognitive decline, and for measuring the ability to adjust these intentions according to knowledge about prevention, risk and symptoms. However, we recognize the need for further investigation. The finding of differential average scores and item functioning between English and Spanish versions warrants exploration. Future research studies should test this instrument in larger and more representative samples. This expanded testing should include participants from various Latino subgroups, considering factors such as country of origin, acculturation level, and socioeconomic status. Such comprehensive evaluation will help to identify any potential biases or limitations in the instrument’s applicability across different Latino communities and will help to maximize its accuracy and utility in both research and clinical practice.

## Data Availability

All data produced in the present study are available upon reasonable request to the authors.

## Acknowledgement

We sincerely thank the other contributors, without whom this work would not have been possible. These invaluable individuals include Diana Martinez Garcia and Melanie Benito, for their invaluable assistance in developing the instrument and recruitment efforts; The remarkable team at the University of Wisconsin Survey Center for their work in the planning and development stages of our work; and Uriel Paniagua, Valentina Flores Diaz, Javier Neira Salazar, George Levy, and Maria del Carmen Rosales for their outstanding support throughout this process.

